# Assessing the influence of climate on future wintertime SARS-CoV-2 outbreaks

**DOI:** 10.1101/2020.09.08.20190918

**Authors:** Rachel E. Baker, Wenchang Yang, Gabriel A. Vecchi, C. Jessica E. Metcalf, Bryan T. Grenfell

## Abstract

High susceptibility has limited the role of the climate in the SARS-CoV-2 pandemic to date. However, understanding a possible future effect of climate, as susceptibility declines and the northern-hemisphere winter approaches, is an important open question. Here we use an epidemiological model, constrained by observations, to assess the sensitivity of future SARS-CoV-2 disease trajectories to local climate conditions. We find this sensitivity depends on both the susceptibility of the population and the efficacy of non-pharmaceutical controls (NPIs) in reducing transmission. Assuming high susceptibility, more stringent NPIs may be required to minimize outbreak risk in the winter months. Our results imply a role for meteorological forecasts in projecting outbreak severity, however, reducing uncertainty in epidemiological parameters will likely have a greater impact on generating accurate predictions and reflects the strong leverage of NPIs on future outbreak severity.

---

The SARS-CoV-2 virus has spread across all geographic regions irrespective of local climate. Cases have continued to climb in both hot, humid conditions, such as the southern United States in summer and India during the southwest monsoon, as well as cold, dry conditions such as Wuhan province in China in the winter. High susceptibility has likely limited the role of climate in the early pandemic such that the signature of seasonality is not yet visible [1]. However, as susceptibility starts to decline, particularly in regions with high numbers of cases, the extent to which the climate may determine the future pandemic trajectory remains unclear.

Many directly-transmitted infectious diseases display seasonal cycles of incidence [2]. For several viral infections, including varicella, influenza and respiratory syncytial virus, these cycles have been shown to be dependent on climate [3, 4, 5, 6, 7] (along with seasonal population aggregation such as school terms). Endemic coronaviruses have also demonstrated sensitivity to the climate in both laboratory studies [8] and at the population level [7]. While the novel coronavirus, SARS-CoV-2, also appears to be sensitive to the climate in laboratory settings [9], case data has yet to reveal a clear environmentally-driven trend [10].

In a recent study, we used an epidemiological model to explore the subtleties of pandemic seasonality [1]. We showed that the climate plays a secondary role compared to high susceptibility in determining early pandemic trajectories, yet we identified potential climate impacts as susceptibility waned. Thus, a more detailed investigation of possible (northern hemisphere) wintertime effects is important, especially given the major, locally-variable impacts of NPIs. Here we consider how climate conditions in the coming months may influence the future trajectory of the pandemic as susceptibility declines. We probe the possible effect of climate while varying two factors: the level of depletion of susceptibles and the relative efficacy of non-pharmaceutical interventions (NPIs) in reducing transmission. Building on our prior work [1], we use a climate-driven Susceptible-Infected-Recovered-Susceptible (SIRS) model to simulate the disease dynamics under these different scenarios and across different climates. The model is based on the estimated climate sensitivity of endemic betacoronavirus HKU1 (see Discussion for the effect of varying climate sensitivity). A key question is the extent to which the upcoming northern hemisphere winter climate may exacerbate future cases numbers. To address this, we first consider possible case trajectories for New York City (results for select other northern hemisphere locations are shown in Fig. S1–3).

## Results

In Fig. 1a we use case data (see Methods) to estimate the effective reproductive number of infection for New York City from the start of 2020 to the present (July 2020) [11]. Estimated values of *R_effective_* peak early in the outbreak and then settle close to 1 in the summer months as NPIs act to lower transmission. We assume the *R_effective_* values approximate *R*_0_ and compare them to the predicted seasonal *R*_0_, derived from our climate-driven SIRS model. Current rates (average over second and third weeks of July) of *R_effective_* in New York city are found to be approximately 35% below the *R*_0_ levels predicted by our climate-driven model. To project future scenarios we assume that *R*_0_ remains at either the current levels (constant) or a relative 35% decrease in our climate-driven *R*_0_ which means *R*_0_ oscillates with specific humidity (Fig. 1a, top plot).

**Figure 1:**
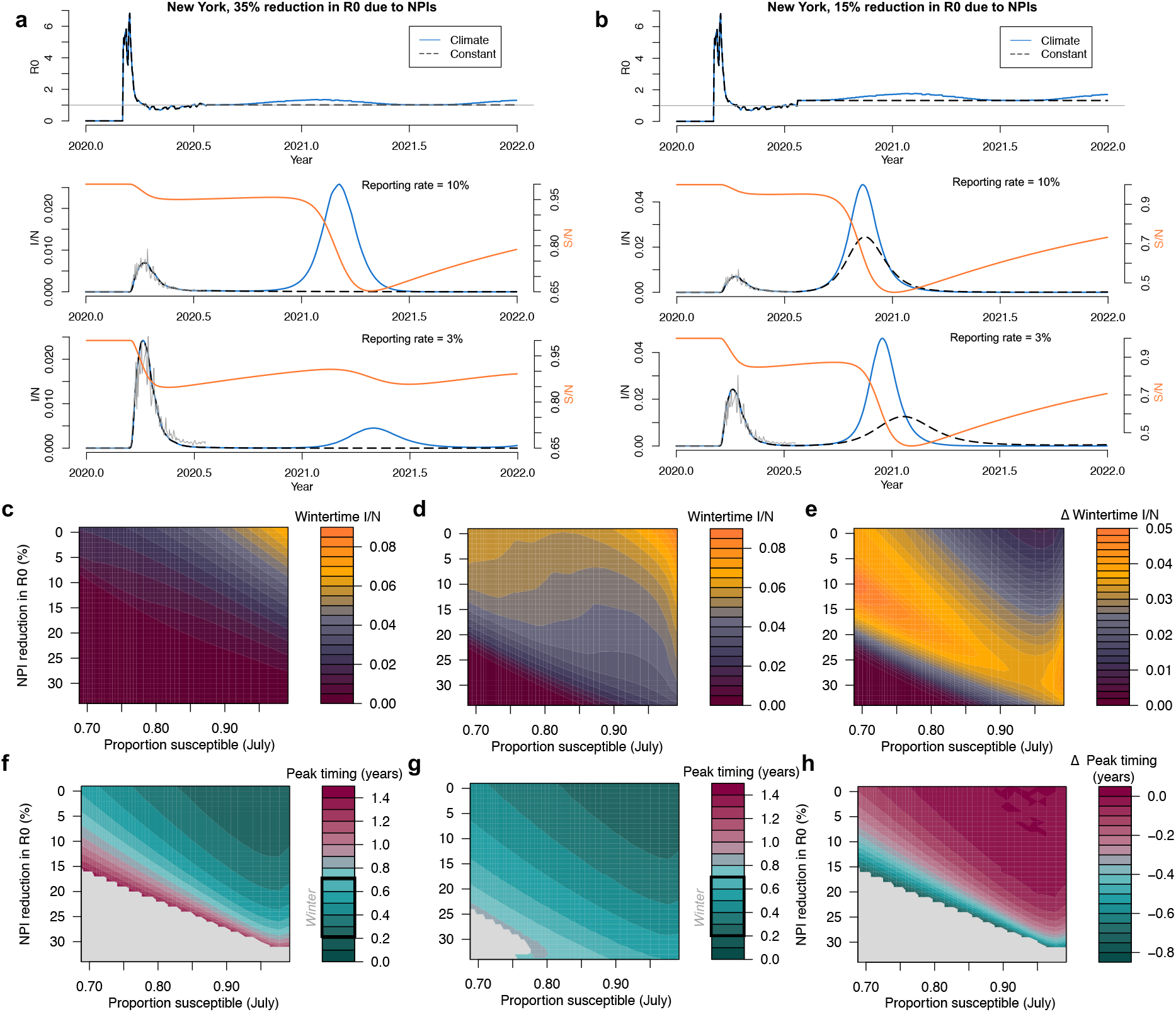
Wintertime outbreaks in New York City. Estimated and projected *R*_0_values (top plot) assuming a) 35% and b) 15% reduction in *R*_0_ due to NPIs. Corresponding time series show the simulated outbreaks in the climate (blue) or constant (black/dashed) scenarios, with middle row plots assuming a 10% reporting rate and bottom row plots assuming a 3% reporting rate. Corresponding susceptible time series are shown in orange. Case data from New York City is shown in grey. Surface plots (top) show the peak wintertime proportion infected (infected = I/ population = N) and (bottom) the timing of peak incidence in years from July in the scenarios with c) the constant *R*_0_ and d) the climate-driven *R*_0_. e) shows the difference between the climate and constant *R*_0_ scenario.

In Fig. 1a (lower plots) we show the proportion infected over time using the climate-driven and constant *R*_0_ values. We also vary the reporting rate of observed cases relative to modelled cases; while this accounts for under-reporting it also allows us to vary the proportion susceptible over a feasible range (see Methods). In the middle figure, the reporting rate is 10% (estimates for US reporting rates are < 10% [12]), which implies a relatively small reduction in susceptibility based on case data pre-July. In this case, a small boost to transmission, driven by low specific humidity in the winter months, results in a relatively large secondary outbreak in the climate scenario. In the constant scenario, *R*_0_ stays below 1 and there is no outbreak in the winter months. We also consider a scenario where the reporting rate is 3% (Fig. 1a lower plot). In this case the lower reporting rate means more cases (relative to the observed case counts) and a greater reduction in susceptibility. This results in a smaller wintertime outbreak in the climate scenario.

In Fig. 1b we consider a scenario where NPI measures are relaxed further such that *R*_0_ is reduced 15% below non-control values as of the last week in July. In this case *R*_0_ *>* 1 for both the climate and constant scenario and case numbers begin to grow exponentially. With a 10% reporting rate a large secondary outbreak is observed in both the constant and climate scenarios (Fig. 1b middle plot). With a 3% reporting rate, meaning a larger depletion of susceptibles, the secondary outbreak appears much larger in the climate scenario: this supports the hypothesis that the disease will become more sensitive to climate as the susceptible proportion declines, much like the seasonal endemic diseases.

In Fig. 1 c-h we simulate model outcomes across a broad range of parameter space varying the proportion susceptible (in July) and the reduction in *R*_0_ due to NPIs. The proportion susceptible is varied by initializing the epidemic with different sizes of the infected population (initializing with a large number results in a relatively larger outbreak and initializing with a small number results in a smaller outbreak). We vary this starting number over a feasible range given the case data i.e. such that observed cases never exceed modelled cases or that the reporting rate never drops below 1%. Over this range, the model plausibly tracks the observed case data.

Fig. 1e shows the change in winter peak size (max proportion infected between September - March) due to climate. Peak size results for the constant and climate scenarios are shown in Fig. 1c and d respectively. When the susceptible proportion is high and the effect of NPIs are minimal (relative *R*_0_ given NPI = 1), large outbreaks are possible in both the climate and constant *R*_0_ scenarios meaning the relative effect of climate on peak size and timing is close to 0 (top right Fig. 1e). As the proportion susceptible declines (moving left along the x-axis of Fig. 1e), case trajectories become more sensitive to the wintertime weather resulting in larger peaks in the climate scenario. However, sufficiently strong NPIs, in combination with low susceptibility, reduce incidence to zero in both the climate and control scenarios (bottom left Fig. 1e). NPIs are not as effective at reducing cases when susceptibility is higher (bottom right Fig. 1e).

We also consider the effect of climate on secondary peak timing. Fig. 1f and g shows the peak timing in years (relative to July 2020) in the constant and climate scenarios respectively. In the climate scenario, peak timing for New York is clustered in the winter months (Fig. 1a-b). In the constant *R*_0_ scenario, secondary peaks can occur at a wide range of times over the next 1.5 years. As in the peak size results, high susceptibility and limited NPIs reduce the effect of climate and peak timing is matched for both the climate and control scenarios (top right Fig. 1h). Grey areas represent regions where there is no secondary peak in either the climate or control scenario.

We next consider the relative effect of climate on peak size for nine global locations (Fig. 2b). In this case, as opposed to using estimated *R_effective_* values (given case data is not available for several of the global cities), we simulate the epidemic from July 2020 using a fixed number of infecteds and vary the starting proportion of susceptibles (example results from select global locations, using estimated *R_effective_*, are shown in Fig. S1–3). Results from the New York surface in Fig. 2b qualitatively match our tailored simulation in Fig. 1. Locations in the southern hemisphere are expected to be close to their maximum wintertime *R*_0_ values in mid 2020 (Fig. 2a) meaning that secondary peaks in the climate scenario are lower than the constant *R*_0_ scenario for these locations. Tropical locations experience minimal difference in the climate versus constant *R*_0_ scenario given the relatively mild seasonal variations in specific humidity in the tropics. Broadly, the results across hemisphere track the earlier results from New York: high susceptibility and a lack of NPIs lead to a limited role of climate, but an increase in NPI efficacy or a reduction in susceptibility may increase climate effects. This result is more striking in regions with a large seasonality in specific humidity.

**Figure 2:**
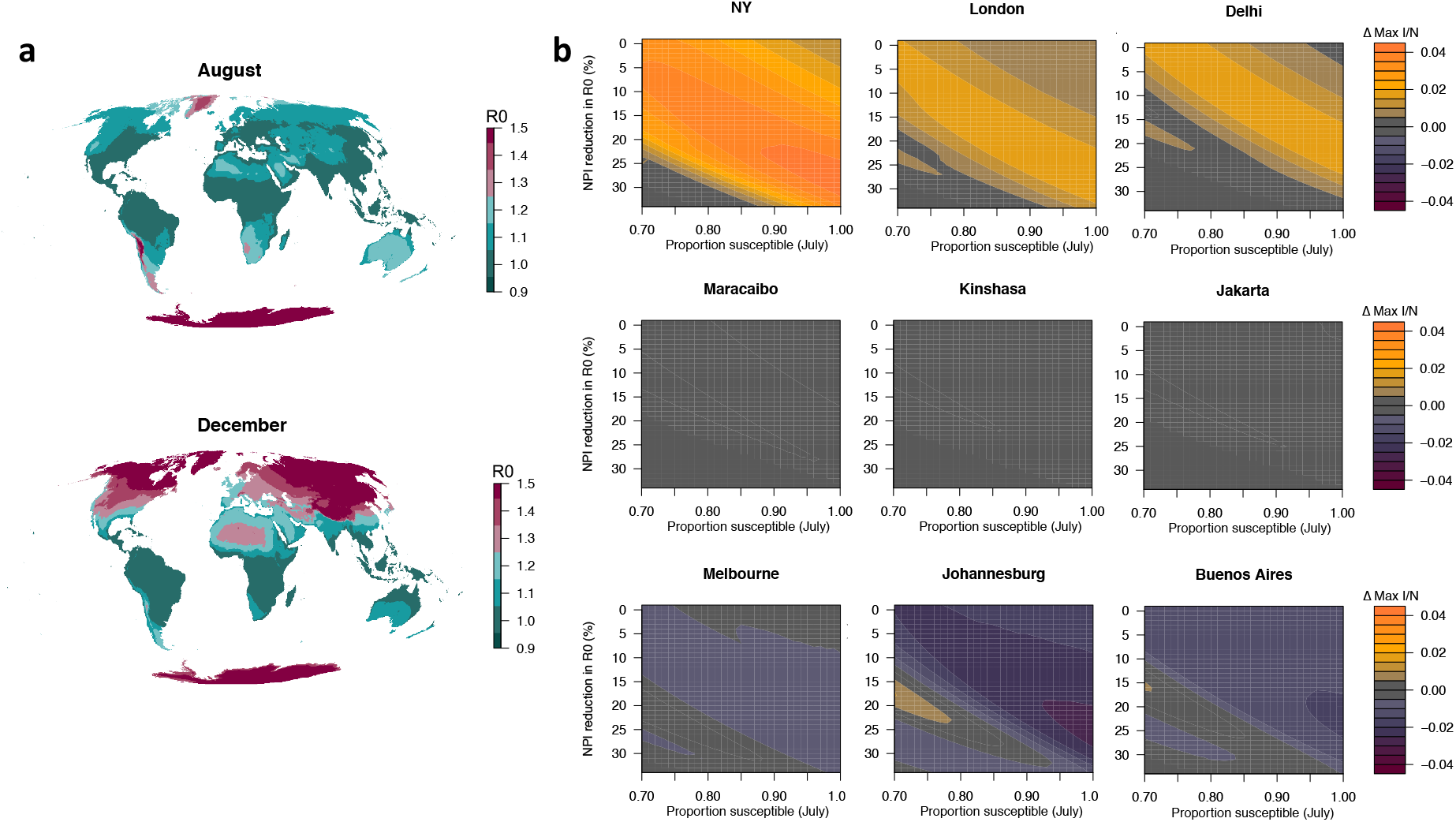
Climate sensitivity of outbreaks across global locations. a) The climate effect on *R*_0_ assuming a 35% reduction due to NPIs shown for August and December. b) The effect of changing susceptibility and NPIs on peak proportion infected (post July 2020) for nine global locations.

Our results suggest that climate may play an increasing role in determining the future course of the SARS-CoV-2 pandemic, depending on levels of susceptibility and NPIs. We next evaluate the extent to which interannual variability in specific humidity could influence peak size. We simulate separate New York pandemic trajectories using 11 years (2008–2018) of specific humidity data. Fig. 3a shows the variability in *R*_0_ and secondary peak size based on these runs (with 35% reduction in *R*_0_ due to NPIs and 10% reporting rate – the same as Fig. 1a). While a relatively large peak occurs in all years, the largest peak (0.038 proportion infected) is almost double the smallest peak year (0.020 proportion infected). In Fig. 3b we calculate the coefficient of variation of the peak size for different susceptible proportions and NPI intensities. These results qualitatively track Fig. 1e. Sensitivity to interannual variation appears most important when the susceptible population has been reduced by at least 20% and minimal controls are in place.

**Figure 3:**
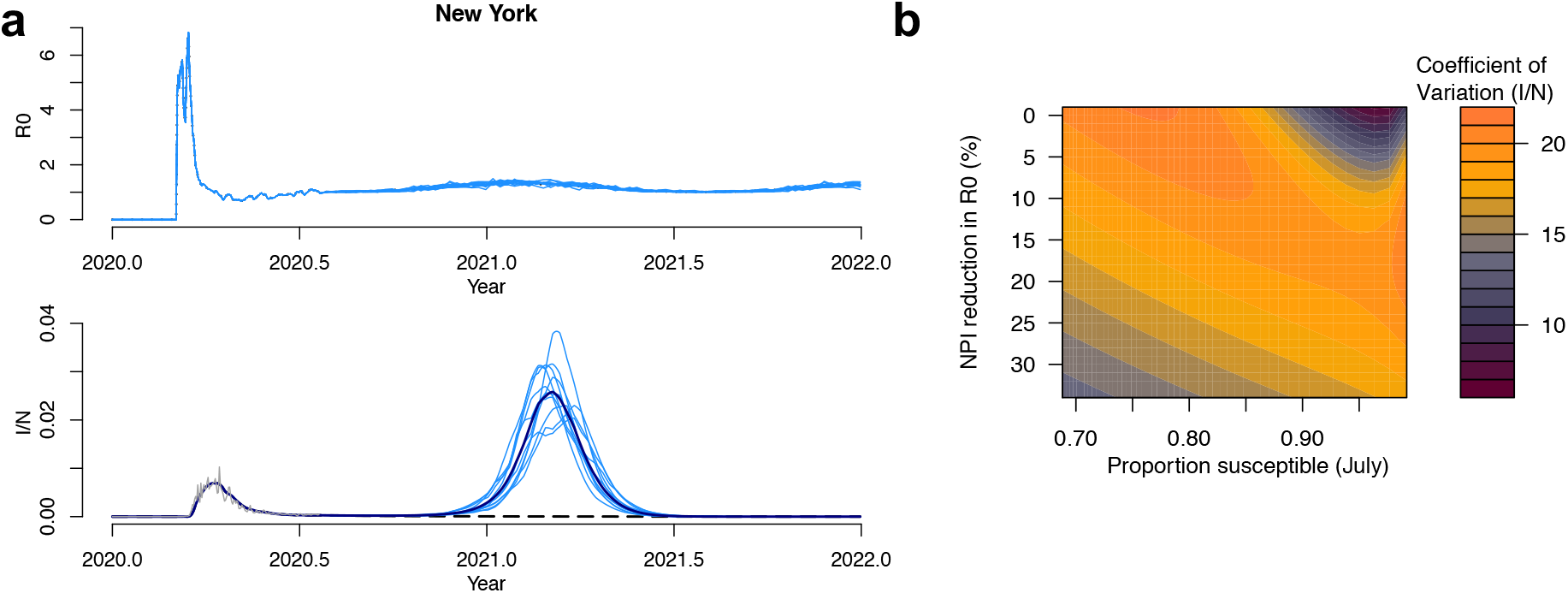
Climate variability and wintertime cases in New York. a) Climate driven *R*_0_ and corresponding infected time series based on the last 10 years of specific humidity data for New York, assuming a 35% reduction due to NPIs. b) The effect of changing susceptibility and NPIs on the coefficient of variation of peak incidence for simulations using specific humidity data from 2008-2018.

## Discussion

There are several caveats to our results. First, the precise mechanism by which climate modulates seasonal transmission rates for viruses is currently unknown. While virus survival and changes to the immune system are expected to fluctuate with the weather, changes to human behavior, such as grouping indoors during cold weather, may partly determine the seasonal effect. Given the broad societal disruptions of the COVID-19 pandemic, these latter behaviors are likely to be reduced, such that total climate-driven fluctuations to transmission may be modified. Further, additional seasonal behaviors that may have also driven transmission, such as population aggregation through schooling, will also be reduced by NPIs.

Second, we do not directly estimate the climate sensitivity of SARS-CoV-2. Studies exploring this relationship using case data have yet to find a conclusive result [10]. Instead our model relies on estimates of the climate-sensitivity of another betacoronavirus, HKU1. We also make an assumption as to the length of immunity to the virus, using estimates for HKU1 (66 weeks) [1]. While the length of immunity may not affect the dynamics in the early stage of the pandemic, it could have complex and uncertain outcomes for future trajectories [13].

We consider the possible contribution of uncertainty in parameters to the variance in the wintertime peak size following the method developed by Yi et al.[14] (see Methods). Fig. 4 shows contribution to variance in wintertime peak size of five parameters: NPIs efficacy, immunity length, reporting rate, climate sensitivity of the virus and interannual climate variability. We find that climate sensitivity is an important factor but secondary to the efficacy of NPIs and immunity length in determining peak transmission. Uncertainty in immunity length and reporting together influence susceptibility and collectively account for the second largest portion of total uncertainty. Uncertainty in interannual variability i.e. climate variability and weather, has a smaller impact on peak size. NPIs contribute the largest proportion to total variance in peak size. It is important to note that while other parameters are external features of either the virus, climate, or disease trajectories to date, the efficacy of NPIs is determined directly by policy interventions and therefore the size of future outbreaks is largely under human control.

**Figure 4:**
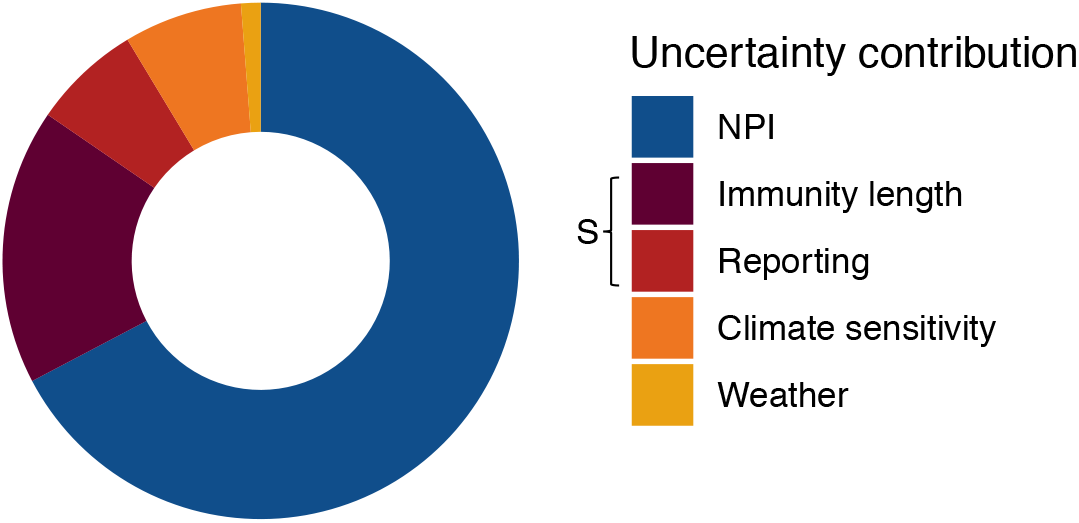
Contribution to uncertainty in New York wintertime 20/21 peak size. The relative importance of NPI efficacy [0 – 35%], immunity length (10 - 60 weeks), reporting (1 - 100%), climate sensitivity of the virus [−32.5 to −227.5] and interannual weather variability [10 years] in determining wintertime peak size. Immunity length and reporting rate collectively determine susceptibility, *S*.

Our results imply that meteorological and climate forecasts could be helpful in predicting future outbreak size (Fig. S4). However, this information will likely be secondary to epidemiological monitoring such as estimates of the efficacy of active control measures in reducing transmission and serological surveys to determine susceptiblity [15]. Current data from serological surveys suggest a minimal reduction in susceptiblity in many locations [16], with New York City towards the upper bound in terms of total reduction in susceptibility [17]. For many other locations, susceptiblity may be much closer to 1, meaning the efficacy of NPIs will be a key determinant of winter outbreak size. Our results therefore suggest that more stringent NPIs may be required during the winter months to minimize total risk.

## Methods

### Data

Global COVID19 case data come from the Johns Hopkins coronavirus resource center. County-level coronavirus cases data, including estimates for New York city, come from the New York Times. Specific humidity data come from ERA5 [18].

### *R*_0_ estimates

We use the EpiEstim package in the R programming language to estimate *R_effective_* from coronavirus cases data assuming an uncertain serial interval with a mean of 4.7 days and a standard deviation of 2.9 days. We calculate *R_effective_* estimates from the first date in 2020 where case numbers are greater than zero for a particular location. *R_effective_* is estimated until the 21st July 2020 (the date we first accessed the data). After 21st July 2020 we use an *R*_0_ value modulated by climate and NPI efficacy. The climate-driven *R*_0_ values are based on the climate-driven SIRS model described previously [1]. Specifically, climate-driven values of *R*_0_ are given by:

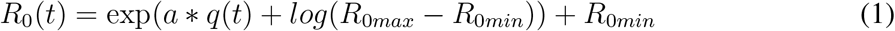

where *R*_0*max*_ and *R*_0*min*_ are the maximum and minimum reproductive numbers respectively, set at 2.5 and 1.5 based on estimates for SARS-CoV-2 [1, 19]. *q*(*t*) is specific humidity and *a* is the climate dependence parameter (set at –227.5) based on model fits for the HKU1 betacoronavirus[1]. We test sensitivty of our results to different values of parameter *a* in Fig. 4.

### SIRS model

Our *R*_0_ estimates are incorporated into an Susceptible-Infected-Recovered-Susceptible model where *R*_0_(*t*) = *β*(*t*)*D*. D is the mean infectious period (set at 5 days) and *β*_(_*t*) is the contact rate. The SIRS model is directly dependent on *β*_(_*t*) and is given by:

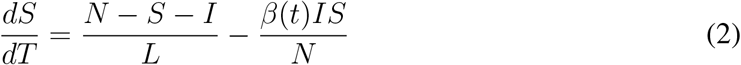

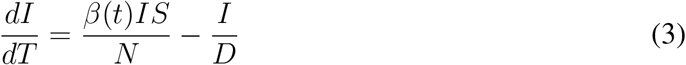

where S is the number of susceptibles, I is the number of infecteds, R is the number if individuals in the recovered category and N is the population size.

We initialize the model on the first day cases are observed. In order to capture different possible trajectories, we initialize varying the proportion infected on the first day. Over a finite range, models initialized with different infected proportions are able to track observed cases to a scaling constant i.e. the reporting rate (Fig. a, b). We tune the range of starting proportion infected such that the reporting rate stays between 1% and 100%, though results for specific trajectories are shown for 3% and 10% in Fig. 1 a-b, reflecting prior estimates on reporting rates [12].

### Uncertainty decomposition

We run our model for New York using ten discrete values of non-pharmaceutical intervention, immunity length, reporting rate (determined by the initial proportion infected), climate sensitivity and weather variability (determined by using historic weather observations from 2009 to 2018): a total of 10,000 model runs. For each model run, we record the wintertime peak size. We then use ANOVA on a fixed effect regression model where the dependent variable is wintertime peak size and the fixed effects are the factorial contribution of each parameter. The total sum of squares is calculated for each parameter across factors. A similar approach has been used to decompose uncertainty in climate model projections [14]. Using fixed effects allows us to recover some of the non-linearity in possible parameter dependence.

To create Fig. 4, we divide the sum of squares attributable to each parameter by the total explained sum of squares. Parameters were varied over a plausible range i.e. NPI efficacy [0 –35%], immunity length [10 – 60 weeks], reporting (1–100%), climate sensitivity (OC43 climate sensitivity of –32.5 to HKU1 climate sensitivity of –227.5 [1]) and weather variability (based on 2009–2018 weather). However, it is important to note that expanding the range of a particular parameter would likely increase the importance of its predicted effect. As such, this method only provides a proxy for considering possible contributions to uncertainty.

## Data Availability

All data is publicly available.

https://github.com/CSSEGISandData/COVID-19

## References

[1] Baker, R. E., Yang, W., Vecchi, G. A., Metcalf, C. J. E. & Grenfell, B. T. Susceptible supply limits the role of climate in the early sars-cov-2 pandemic. Science (2020).

[2] Martinez, M. E. The calendar of epidemics: Seasonal cycles of infectious diseases. PLoS pathogens 14 (2018).

[3] Shaman, J. & Kohn, M. Absolute humidity modulates influenza survival, transmission, and seasonality. Proceedings of the National Academy of Sciences 106, 3243–3248 (2009).

[4] Shaman, J., Pitzer, V. E., Viboud, C., Grenfell, B. T. & Lipsitch, M. Absolute humidity and the seasonal onset of influenza in the continental united states. PLoS biology 8, e1000316 (2010).

[5] Pitzer, V. E. et al. Environmental drivers of the spatiotemporal dynamics of respiratory syncytial virus in the united states. PLoS pathogens 11, e1004591 (2015).

[6] Baker, R. E., Mahmud, A. S. & Metcalf, C. J. E. Dynamic response of airborne infections to climate change: predictions for varicella. Climatic Change 148, 547–560 (2018).

[7] Baker, R. E. et al. Epidemic dynamics of respiratory syncytial virus in current and future climates. Nature Communications 10, 1–8 (2019).

[8] Chan, K.-H. et al. The effects of temperature and relative humidity on the viability of the sars coronavirus. Advances in virology 2011 (2011).

[9] Chin, A. W. et al. Stability of sars-cov-2 in different environmental conditions. The Lancet Microbe 1, e10 (2020).

[10] Smit, A. et al. Winter is coming: A southern hemisphere perspective of the environmental drivers of sars-cov-2 and the potential seasonality of covid-19 (2020).

[11] Wallinga, J. & Teunis, P. Different epidemic curves for severe acute respiratory syndrome reveal similar impacts of control measures. American Journal of epidemiology 160, 509–516 (2004).

[12] Perkins, A. et al. Estimating unobserved sars-cov-2 infections in the united states. PNAS (2020).

[13] Saad-Roy, C. M. et al. Immuno-epidemiological life-history and the dynamics of sarscov-2 over the next five years. medRxiv (2020).

[14] Yip, S., Ferro, C. A., Stephenson, D. B. & Hawkins, E. A simple, coherent framework for partitioning uncertainty in climate predictions. Journal of Climate 24, 4634–4643 (2011).

[15] Arora, R. K. et al. Serotracker: a global sars-cov-2 seroprevalence dashboard. The Lancet. Infectious Diseases (2020).

[16] Havers, F. P. et al. Seroprevalence of antibodies to sars-cov-2 in 10 sites in the united states, march 23-may 12, 2020. JAMA Internal Medicine (2020).

[17] Rosenberg, E. S. et al. Cumulative incidence and diagnosis of sars-cov-2 infection in new york. medRxiv (2020).

[18] Hersbach, H. et al. The era5 global reanalysis. Quarterly Journal of the Royal Meteorological Society (2020).

[19] Kissler, S. M., Tedijanto, C., Goldstein, E., Grad, Y. H. & Lipsitch, M. Projecting the transmission dynamics of sars-cov-2 through the post-pandemic period. Science (2020).

